# Degree of immunoglobulin kappa light chain glycosylation of anti-spike SARS CoV-2 antibodies correlates with COVID-19 severity

**DOI:** 10.1101/2023.01.06.23284259

**Authors:** Raminta Grigaite, Jason K Iles, Stephen Harding, Roshani Patel, Gregg Wallis, Ray K Iles

## Abstract

Glycosylation of antibodies and the effects this has on inflammatory responses has concentrated predominately on the study of glycosylation moieties found in the Fc region of heavy chains. Light chain glycosylation and their ratios are relatively understudied. Nevertheless, variable glycosylation and ratio of κ and λ light chains have been associated with worse prognosis in myeloma and in tissue deposition – amyloidosis.

The κ & λ light chains, of antibodies binding to SARS-CoV2 nucleocapsid and spike protein were analysed, using MALDI-ToF MS, in respect to their intensity, ratios, glycosylation patterns and any pattern changes correlating with COVID-19 severity. The molecular masses and signal intensity of κ and λ glycosylated and non-glycosylated light chains were measured for immunoglobulins isolated from plasma of sero-positive and sero-negative health care workers (HCW), and convalescent patients who had suffered from acute respiratory distress syndrome (ARDS).

Overall, there was no significant changes in κ to λ ratio of total IgG (via protein G capture) antibodies between the groups. A non-statistically significant trend towards λ light chains was found in antibodies against SARS CoV-2 Nucleocapsid and Spike proteins. However, detailed analysis of the molecular forms found a significant increase and bias towards un-glycosylated light chains and in particular un-glycosylated κ light chains, in antibodies against SAR-CoV-2 spike protein, from convalescent COVID-ARDS patients.

Here we have demonstrated a bias towards un-glycosylated κ chains in anti-spike antibodies in those who suffered from ARDS as a result of SARS-CoV2 infection 3 months after recovery. How this relates to the immunopathology of COVID-19 requires further study.

## Introduction

A novel zoonotic coronavirus (SARS-CoV2) was first reported in December 2019 [1]. It since has caused a worldwide pandemic of COVID-19 disease with over 636 million reported cases and over 6,600,000 lives lost [2]. There has been a tremendous effort by scientists worldwide researching the testing, vaccine development, risk factors, new treatments and more, with PubMed database giving over 170,000 results on COVID-19. Our group has focused on the SARS-CoV2 Spike and Nucleocapsid protein binding of serum proteins [3], [4] and direct detection of humoral markers for rapid and direct population screening. The main focus was the identification of those most at risk, by direct MALDI-ToF mass spectrometry analysis of human serum albumin and immunoglobulin heavy chains [5].

Two isotypes of polyclonal light chains, κ and λ, are synthesised in plasma cells that are then matched to the heavy chain (α, δ, ε, γ and μ) by the disulphide bonds to form a functioning immunoglobulins (IgA, IgD, IgE, IgG and IgM, respectively) [6]. Following immunoglobin expression further post-translational modifications take place. Glycosylation is one of the key regulators of antibody’s biological activity, controlling antibody’s stability, half-life, secretion, immunogenicity and function [7]. Variable pattern of type and amount of glycosylation in Fab region of antibody is known to play a role in pathophysiology, particularly in autoimmunity and oncology [8] and with more data the role of glycosylation is thought to be of critical importance [9], [10]. Unlike in autoimmune and oncologic conditions, the ability to capture pathogen-specific, in this case SARS-CoV-2 specific, antibodies give us an opportunity to observe disease dependent changes.

As part of the investigation, we have observed the variability of immunoglobulin light chain (Ig Lc) κ and λ ratios and their glycosylation patterns. The molecular masses of κ and λ glycosylated light chains have been confirmed by Kumar et al., [11]. Researchers have used monoclonal antibodies to capture and isolate free κ and λ light chains, before mass spectral analysis by MALDI-ToF MS. Following by PNGase F digestion of the isolate κ and λ chains the mass differences in the immunocaptured forms were confirmed to be due to N-linked glycosylation of the free light chains [11]. We had noted up to four resolving mass forms of light chain in our capture of anti-SARS-CoV-2 antibodies.

In this study we present our findings on the magnetic bead conjugated with immobilised SARS-CoV2 Spike and Nucleocapsid proteins captured immunoglobulin’s light chains. Molecular signature intensity and molecular weight of bound proteins were recorded and relatively quantified by MALDI-ToF mass spectrometry. The particular focus was given to the antibody κ and λ light chains and their glycosylated variants. The ratios and mass shifts of the glycosylated forms were analysed in relation to the COVID-19 severity by analysis of convalescent plasma.

## Material and Methods

### Samples

Serum and plasma samples were obtained from Health care workers (HCWs) and patients referred to the Royal Papworth Hospital for critical care. COVID-19 patients hospitalised during the first wave and as well as NHS healthcare workers working at the Royal Papworth Hospital in Cambridge, UK served as the exposed HCW cohort (Study approved by Research Ethics Committee Wales, IRAS: 96194 12/WA/0148. Amendment 5). NHS HCW participants from the Royal Papworth Hospital were recruited through staff email over the course of 2 months (20^th^ April 2020-10^th^ June 2020) as part of a prospective study to establish seroprevalence and immune correlates of protective immunity to SARS-CoV-2. Patients were recruited in convalescence either pre-discharge or at the first post-discharge clinical review. All participants provided written, informed consent prior to enrolment in the study. Sera from NHS HCW and patients were collected between July and September 2020, approximately 3 months after they were enrolled in the study.

For cross-sectional comparison, representative convalescent serum and plasma samples from seronegative HCWs, seropositive HCW and convalescent PCR-positive COVID-19 patients were obtained. The serological screening to classify convalescent HCW as positive or negative was done according to the results provided by a CE-validated Luminex assay detecting N-, RBD- and S-specific IgG, a lateral flow diagnostic test (IgG/IgM) and an Electro-chemiluminescence assay (ECLIA) detecting N- and S-specific IgG. Any sample that produced a positive result by any of these assays was classified as positive. Thus, the panel of convalescent plasma samples (3 months post-infection) were grouped in three categories: A) Seronegative HCWs (N = 30 samples) B) Seropositive HCWs (N = 31 samples); C) COVID-19 Patients (N = 38 samples) [12].

### Antigen coupled magnetic beads

Protein-G coupled magnetic beads were purchased from Cytivia Ltd (Amersham Place, Little Chalfont, Buckinghamshire, UK). Recombinant nucleocapsid and recombinant stabilized complete spike protein magnetic beads were made by Bindingsite Ltd (Birmingham, UK).

The viral spike protein (S-protein) is present on virions as pre-fusion trimers with the receptor binding domain of the S1 region stochastically open or closed, an intermediary where the S1 region is cleaved and discarded, and the S2 undergoes major confirmation changes to expose and then retract its fusion peptide domain [13]. Here the S-protein was modified to disable the S1/S2 cleavage site and maintain the pre-fusion stochastic confirmation [14].

### Semi-automated magnetic bead capture processing

The processes of magnetic bead capture, washing, agitation and target binding protein elution can vary dramatically due to damage from too vigorous mixing and yet insufficient washing can result in large amounts of non-specific binding proteins being recovered. To minimise these problems, and individual operator variability, in the efficiency of target binding proteins recoveries, the Crick automated magnetic rack system was employed. This has been described in full previously [4].

### Pre-processing of the magnetic beads

1.5µl microcentrifuge tubes were loaded into the automated magnetic rack. Protein G (GE), purified nucleocapsid or purified stabilized complete spike magnetic beads (Bindingsite, Birmingham UK) in their buffer solutions were vortexed to ensure an even distribution of beads within the solution. 10µl of the appropriate magnetic beads were pipetted into each tube. 100µl of wash buffer, 0.1% Tween 20 in Dulbecco’s phosphate, buffered saline (DPBS) was pipetted into each tube before resuspending. After mixing for several minutes, the instrument pulled the antigen beads to one side allowing the wash buffer to be carefully discarded. The wash cycle was repeated three times.

### Sample processing and binding fraction elution from the magnetic beads

45µl of 10X DPBS was pipetted into each of the tubes containing the washed magnetic beads. 5µl of vortexed neat plasma was pipetted and pump mixed into a tube containing the beads, repeating for each plasma sample. After the resuspension, and automated mixing for 20 minutes, the beads were magnetically collected to one side and the non-bound sample discarded. A further 3 wash cycles were conducted using 0.1% DPBS. Subsequently, another 3 wash cycles were conducted after this, using ultra-pure water, discarding the water after the last cycle. 15µl of recovery solution (20mM tris(2-carboxyethyl)phosphine (TCEP) (SigmaAldrich, UK) + 5% acetic acid + ultra-pure water) was pipetted into the tubes. The tubes were run alternatively between the ‘Resuspend’ and ‘Mix’ setting for several minutes. After pulling the extracted magnetic beads to one side the recovery solution was carefully removed using a pipette and placed into a clean, labelled 0.6µl microcentrifuge tube. This recovery solution was the eluant from the beads and contained the desired proteins.

### Sample Analysis by MALDI-ToF Mass spectrometry

Mass spectra were generated using a 15mg/ml concentration of sinapinic acid (SA) matrix. The elute from the beads was used to plate with no further processing. 1µl of the eluted samples were taken and plated on a 96 well stainless-steel target plate using a sandwich technique. The MALDI-ToF mass spectrometer (microflex® LT/SH, Bruker, Coventry, UK) was calibrated using a 2-point calibration of 2mg/ml bovine serum albumin (33,200 m/z and 66,400 m/z) (PierceTM, ThermoFisher Scientific). Mass spectral data were generated in a positive linear mode. The laser power was set at 65% and the spectra was generated at a mass range between 10,000 to 200,000 m/z; pulsed extraction set to 1400ns. A square raster pattern consisting of 15 shots and 500 positions per sample was used to give 7500 total profiles per sample. An average of these profiles was generated for each sample, giving a reliable and accurate representation of the sample across the well. The raw, averaged spectral data was then exported in a text file format to undergo further mathematical analysis.

### Spectral Data processing

Mass spectral data generated by the MALDI-ToF instrument were uploaded to an open-source mass spectrometry analysis software mMass™ [15], where it was processed by using; a single cycle, Gaussian smoothing method with a window size of 300 m/z, and baseline correction with applicable precision and relative offset depending on the baseline of each individual spectra. In the software, an automated peak-picking was applied to produce peak list which was then tabulated and used in subsequent statistical analysis.

### Statistical analysis

Peak mass and peak intensities were tabulated in excel and plotted in graphic comparisons of distributions for each antigen capture and patient sample group. Means and Medians were calculated and, given the asymmetric distributions found, non-parametric statistics were applied, such as Chi^2^ and Kruskal Wallis test, when comparing differences in group distributions.

## Results

This study has utilised the previously described antigen-coupled magnetic bead capture method [4]. Post elution from protein G, and SARS-CoV-2 antigens; S and N, coupled magnetic beads, MALDI-ToF mass spectra was obtained and peaks recorded. Sample treatment with TCEP effectively reduced the disulphide bonds between light and heavy chains of the immunoglobins, enabling mass peaks to be recorded (Figure 1). Molecular weight expressed as m/z as well as peak intensities of Ig κ and λ light chains (∼23,200 m/z, ∼22,600 m/z) and the respective glycosylated forms (∼24,900 m/z, ∼23,800 m/z) were obtained for each antigen binding.

**Figure 1:**
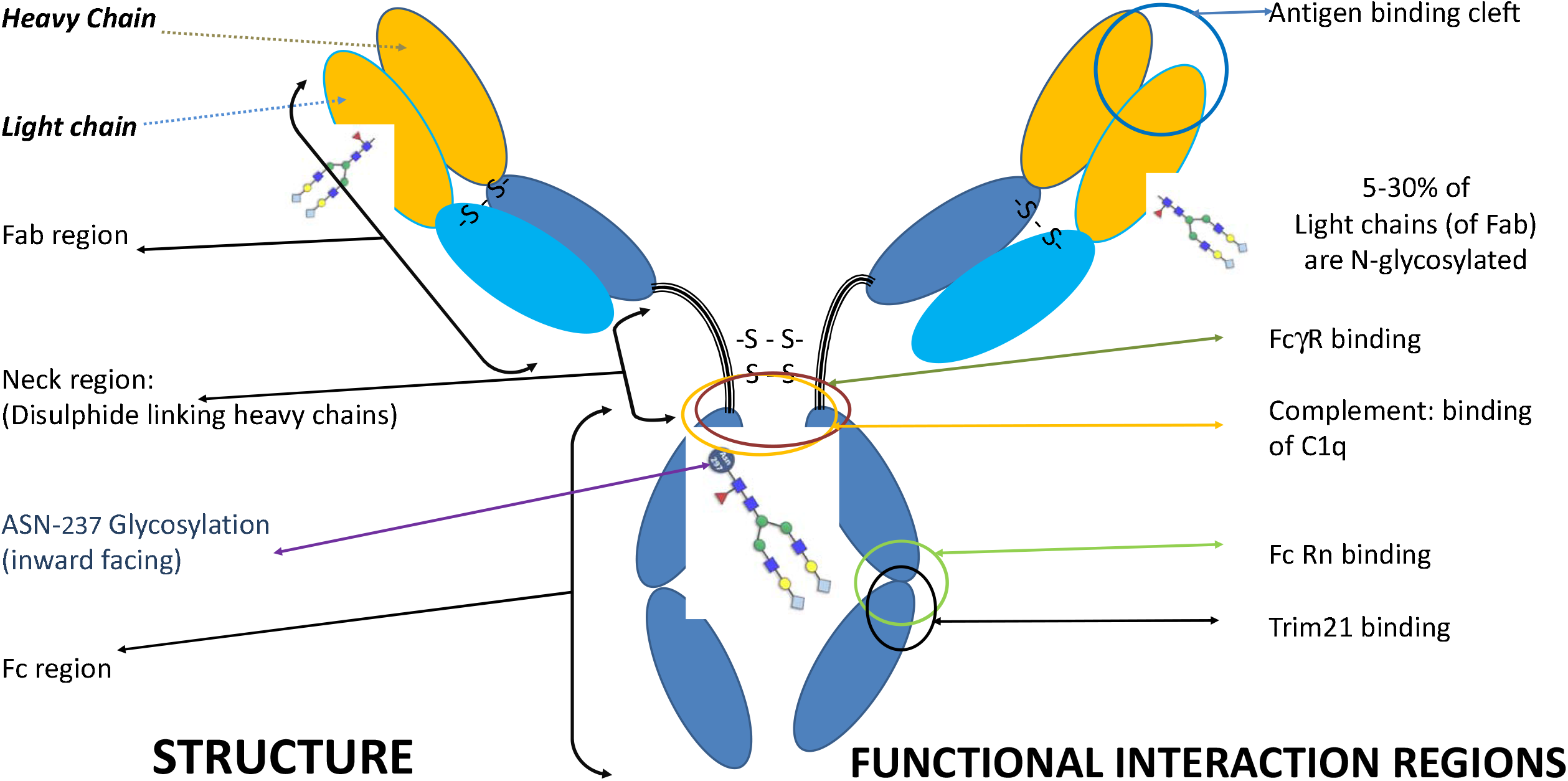
Diagrammatic representation of IgG1 illustrating structure domains and functional mapped regions in relationship to heavy and light chains and glycosylation.

### Immunoglobulin light chain detection

Light chains κ and λ and their glycosylated forms were not detected in all sample outcome groups. Detection proportion also varied depending on the antigen. Most detection was seen with protein G coupled beads capture of sample, non-specific, IgG. High concentrations of un-glycosylated κ & λ, glycosylated λ were detected in all samples examined. However glycosylated κ could not be visualised as it was masked by abundant doubly charged IgG1 heavy chain in this set of (total IgG) isolation experiments. (see Figure 2, and Figure 3). The intensity of Immunoglobulin light chain peaks seen in SAR-CoV-2 antibody capture preparations were on average a log10 lower that the total IgG (protein-G captured) antibody controls (see Figure 4). Anti-nucleocapsid had the lowest capture, overall ranging from 30% to 70% of all samples displaying the various light chain mass peaks, and anti-protein S bound light chain detection was from 48% to 92%, depending on the light chain isotype. In most cases the least detected was κ light chains and the most glycosylated-λ. However, in the anti-S-protein bound light chains of the COVID-19 patient group. un-glycosylated κ light chains were significantly higher and more frequently detected in the plasma samples (p<0.001) (Figure 4).

**Figure 2.**
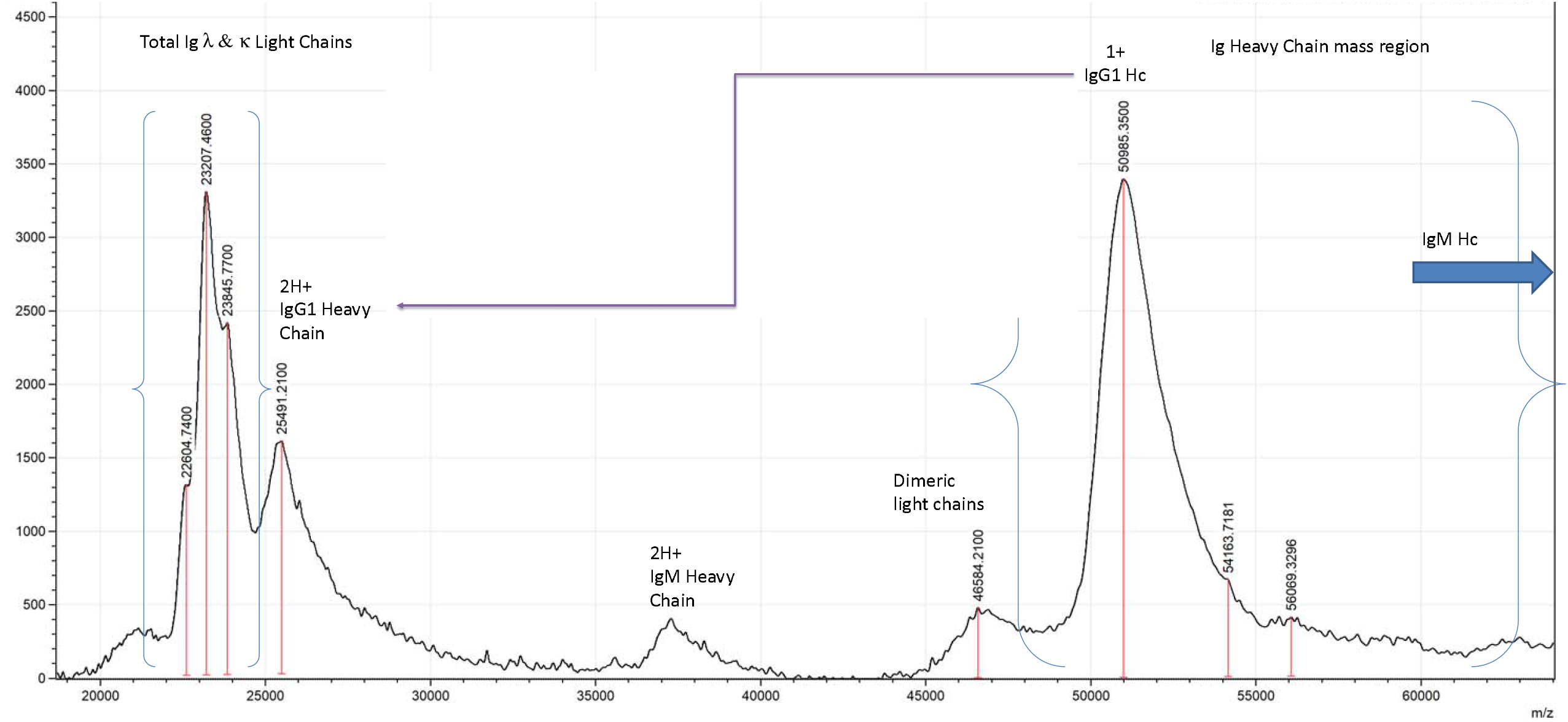
Example Mass spectra profile, (18,000m/z to 64,000m/z), of protein-G coupled magnetic bead captured total IgG from human plasma (eluted at pH 2 and treated with TCEP) showing light and heavy chains of immunoglobulins.

**Figure 3.**
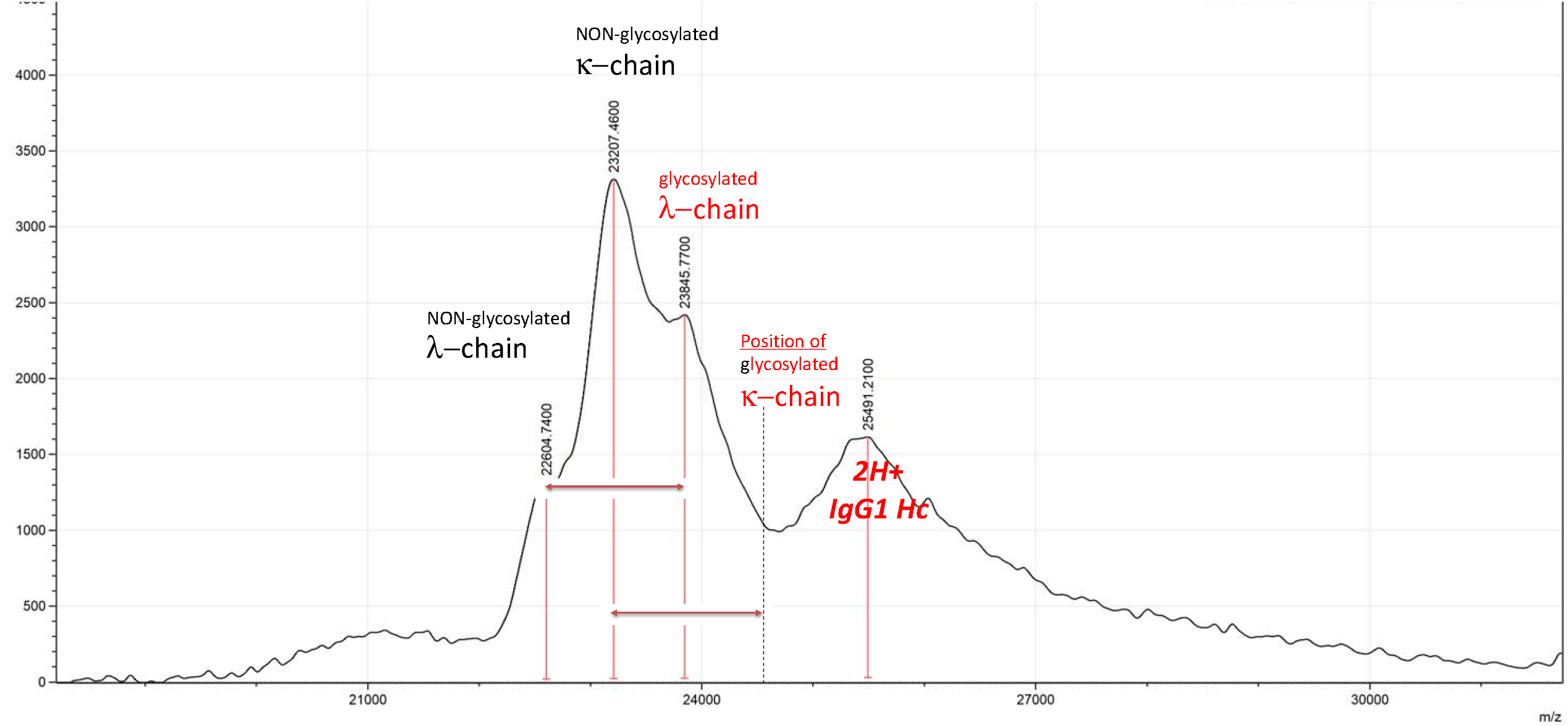
Mass spectra profile zooming to 18,000m/z to 31,000m/z in order to reveal κ and λ light chains (∼23,200 m/z, ∼22,600 m/z) and the respective glycosylated forms (∼25,500 m/z, ∼23,850 m/z).

**Figure 4.**
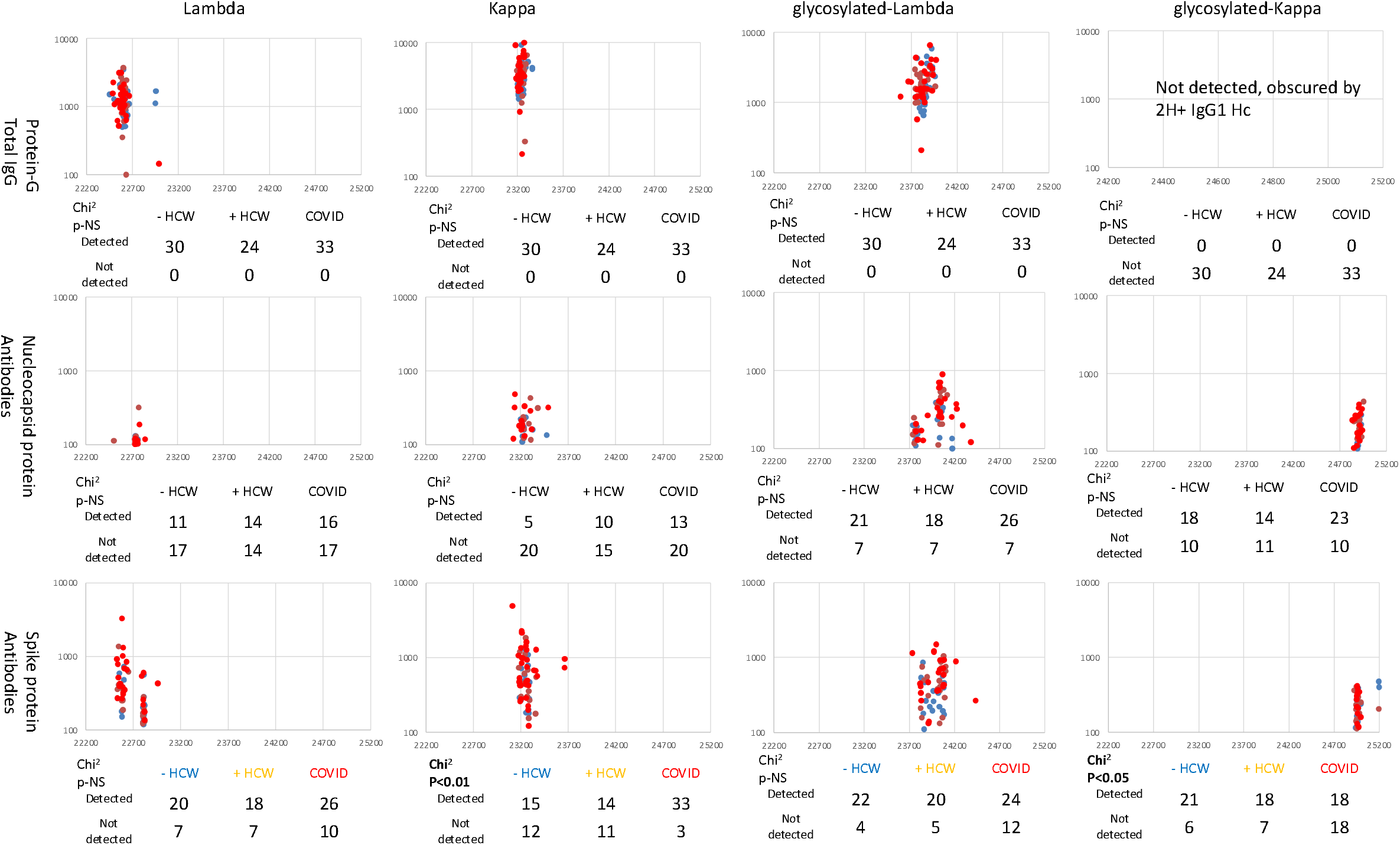
The relative number and distribution of m/z for captured and eluted antibody λ, κ, glc-λ and glc-κ, light chains. Data represents capture antibodies from protein G (total IgG), anti SARS-CoV2 Nucleocapsid and Spike protein, for the different sample groups: Blue represents data from SARS-CoV2 sero-negative HCW, Orange from SARS-CoV2 HCW sero-positive having recovered from COVID-19 with mild symptoms and Red sample data from convalescent patients recovering from COVID-19 ARDS.

### Molecular mass and intensity distribution

Molecular mass and relative peak intensity were measured for all detected immunoglobulin light chains and their glycosylated variants. The m/z values representing molecular weight for both non-glycosylated κ and λ light chains were in a relatively narrow range, showing little variation. However, the m/z values for glycosylated forms of κ and λ light chains have shown some patterns (see Figure 4). The glycosylated λ light chain m/z variation was comparable across G-protein, SARS-CoV-2 S and N protein coupled bead extractions, although more spread (5^th^ and 95^th^ percentile 23,747 m/z and 24,174 m/z, Δ 427 m/z) when compared to non-glycosylated form (5^th^ and 95^th^ percentile 22,510 m/z and 22,817 m/z, Δ 307 m/z). The non-glycosylated κ light chain mass varied the least (5^th^ and 95^th^ percentile 23,180 m/z and 23,380 m/z, Δ 200 m/z) while glycosylated κ-chain varied the most (5^th^ and 95^th^ percentile 24,800 m/z and 25,200 m/z, Δ 400 m/z). Furthermore, distinguishable groups of glycosylated κ-chains were observed clustering around two m/z values (Figure 4). For anti-N-protein antibodies the majority of the detected glycosylated-κ Lc mass was detected at 24,896 m/z. Anti S-protein antibodies had two distinct masses of glycosylated-κ Lc, with values clustering around the medians of 24,955 m/z, and 25,225 m/z.

### Light chain ratio analysis

The observed median κ/λ ratio among all the patient groups across the antibodies captured were within a normal range (reported normal range light chains κ/λ ratio is 0.26 – 1.65). However, the variation in κ/λ ratios varied wildly for the individual patients anti-SARS-Cov-2 antibodies. Both glycosylated Lc and non-glycosylated Lc were summed and compared, median and mean values were reported (Table 1).

**Table.** Ratios of κ and λ light chain and percentage glycosylation of the respective light chains. Data represents captured antibodies from protein G (total IgGs), anti SARS-CoV2 Nucleocapsid and Spike protein antibodies, for the different sample groups: sero-negative HCW, sero-positive having recovered from mild symptoms HCW and convalescent patients recovering from COVID-19 ARDS.

| Isolated Antibodies |  | Sero – negative HCW<br>Median/Mean | Sero-positive HCW<br>Median/Mean | COVID-19 ARDS PATIENTS<br>Median/Mean |
| --- | --- | --- | --- | --- |
| **Protein–G**<br>**Total IgG** | κ/λ ratio | 1.16/1.13 | 1.04/1.14 | 1.10/1.13 |
|  | % glycosylated Light Chains | 27.5/28.4 | 26.0/26.0 | 27.0/27.6 |
|  | % λ glycosylated | 61.4/60.4 | 52.1/53.7 | 58.1/58.7 |
|  | % κ glycosylated | Not detected, obscured by 2H+ IgG1 Hc mass peak |  |  |
| **Anti SARS- COV-2 Nucleocapsid** | κ/λ ratio | 0.67/14.58 | 0.72/20.9 | 0.78/16.0 |
|  | % glycosylated Light Chains | 64/54.2 | 76/63.3 | 70/64.2 |
|  | % λ glycosylated | 51.4/57.7 | 77.6/71.9 | 81.4/73.4 |
|  | % κ glycosylated | 60/62.6 | 53.4/59.5 | 73/59.8 |
| **Anti SARS- COV-2 Spike** | κ/λ ratio | 0.97/1.19 | 0.85/9.8 | 1.03/11.2 |
|  | % glycosylated Light Chains | 54/54.3 | 49/53.2 | 32/35.2* |
|  | % λ glycosylated | 60.8/58.6 | 55/58.4 | 44.7/59 |
|  | % κ glycosylated | 41.1/52 | 44.7/51 | 5.9/22.7** |

Kruskal Wallis test \* p<0.05 \*\* p<0.001

Of the light chains in the total IgG fraction, caught by protein G, about 27% where glycosylated regardless of patient group and just over 50% of λ light chains were glycosylated. Of the anti-SARS-CoV-2 nucleocapsid antibodies between 50 and 80% were glycosylated and this was regardless of being κ or λ. Of the anti-SARS-CoV2 spike protein antibodies from the HCW worker groups, approx. 50% of the light chains were glycosylated. However, in the COVID convalescent patients only about 32% are glycosylated (p<0.05). Of these, HCWs λ light chains averaged 55-60% glycosylated and COVID-19 convalescent patient 45-60% glycosylated (no significant difference); but whilst 55-60% κ light chains of the HCWs were glycosylated, only 6-23% of COVID-19 patients were (p<0.001) (see Table 1).

## Discussion

Antibodies mediate many functions, including neutralisation, cytotoxicity, cytokine release and phagocytosis, playing a key role in protecting against many microbial diseases, including viruses. Aside from antigen recognition and neutralisation specific antibody’s biological activity is regulated by two key factors: genetic selection of the heavy chains and variable glycosylation [16]. However, the vast majority of studies concerning glycosylation altering inflammatory function has focussed on heavy chain glycans below the hinge and within the Fc domains [17], [18]. Nevertheless, Fab region glycosylation has been postulated to be detrimental to the course of infection through decreased antigen binding and reduced neutralisation. Light chain glycosylation also plays role in Ig aggregation and immune complex formation, where such Lc are more prone to amyloidogenicity [8]. It was reported that glycosylation of light chains is four-fold higher in multiple myeloma (MM) patients with amyloidosis compared to MM patients without amyloidosis [19]. Crystal-globulin formation is also associated with abnormal glycosylation, specifically the modification of N-glycans mostly on κ light chains of IgG [20]. In cold agglutinin disease (CAD) terminal sialic acid residues provide additional electrostatic contacts, leading to precipitation [21]. More recently, it was reported that detection of increased glycosylation of light chains could be a useful tool helping to identify patients more at risk from progression to amyloidosis and other plasma cell disorders [11], [22], [23].

These studies illustrate the importance of immunoglobulin Fab region glycosylation patterns and particularly of the Light chain component. It was previously demonstrated, that N-glycosylation sites on the Fab region are acquired following the somatic hypermutations and are present in about 15% of IgGs [24]. Variable glycosylation allows for flexible and specific optimisation of antibodies. It is also controlled by epigenetics, responding to environmental factors [25].

When it comes to glycoprotein analysis, MALDI-ToF mass spectrometry, being a sensitive high throughput technique with a high signal to noise ratio and resolution; has improved the performance for higher mass glycoprotein analysis [26]. Precise mass change identification of the glycoproteins is possible, with changes in mass correlating to known glycans. Glycation and/or glycosylation of the protein increases the mass to charge (m/z), a proxy for molecular weight [27]. This can be as much as by 1.4 kDa to 2.4 kDa per for each N-linked glycan and 300 Da to 1,000 da per each O-linked glycan [26]. However, O-linked glycosylation has not been reported in immunoglobulin light chains; except in recombinant, therapeutic, antibodies engineered in order to alter their pharmacokinetic/pharmacodynamics properties [8], [28]. A distinct glycosylation profile of anti-SARS-CoV 2 IgG was previously reported, with changes associated with increasing COVID-19 disease severity [29]. Here we report the mass analysis of captured SARS-CoV2 specific, Ig light chains to observe disease dependent patterns. In this study, the average mass increase for glycosylated κ lc *was 1,600 m/z* and for λ light chain – 1,250 m/z, when compared to the non-glycosylated forms. We have also shown variable glycosylation, particularly of κ Light chains in patients with antibodies against SARS-CoV-2 antigens (Figure 3 and table 1).

SARS-CoV-2 Spike and Nucleocapsid antigens were both bound by glycosylated light chains, suggesting light chain glycosylation does not hinder antigen recognition and therefore neutralisation in the case of Spike protein antibodies. However, based on current research, light chain glycosylation may inhibit recruitment of other immune cells and even complement [16]. Thus, a preponderance of un-glycosylated light chain antibodies, particularly κ light chains with their propensity to dimerise [11], would be consistent with an exaggerated immune response, excessive cytokine release and effector cell activation leading to non-specific tissue damage characteristic of ARDS [30].

The data presented here indicate a lack of glycosylation of light chains is associated with disease severity in recovering ARDS patients. This is in contrast to heavy chains, where it was reported that increased glycosylated of IgG3 is associated with the severity of COVID-19 disease [5]. Although conversely, afucosylation [31] and decrease in both bisecting N-acetylglucosamine and galactosylation of “IgG” (subclass unspecified) was reported to be associated with COVID-19 disease severity [32]. It is important to note that the above finding was only observed for SARS-CoV-2 Spike protein captured antibody light chains. There was far more glycosylated Lc captured antibodies by SARS-Cov-2 Nucleocapsid protein. The difference of glycosylated Lc proportion between the SARS-CoV2 antibodies against Spike and Nucleocapsid antigens invites further research.

An important disadvantage of this study, however, is that plasma used for COVID-19 ARDS patients was convalescent and the opportunity to observe changes if the longitudinal data, including from acute disease phase, was missed. Nevertheless, since light chain glycosylation and their ratios are relatively understudied, we have presented further insight into κ and λ ratios and their glycosylation patterns. Future studies should consider the functional significance of light chains modifications in respect of COVID-19 and other infectious disease immunopathology.

## Data Availability

All data produced in the present study are available upon reasonable request to the authors

## Abbreviations

HCW: Health care workers
ARDS: Acute respiratory distress syndrome
ITU: Intensive Therapy unit
AGE: advanced glycation end products
MALDI-ToF: Matrix assisted laser desorption ionization – time of flight.

## Acknowledgements

We thanks Dr Helen Baxendale of the Humoral Immune Correlates to COVID-19 (HICC) consortium, funded by the UKRI and NIHR; grant number G107217 (COV0170 - HICC: Humoral Immune Correlates for COVID-19) for providing samples and part funding the study. We also thank the Royal Papworth Hospital Foundation Trust COVID-19 Research and Clinical teams including Intensive care clinicians, Catherine Denny, Maria Nizami, Erin Hopley for supporting recruitment to this study and patients and HCWs who participated in this study.

We grateful thank Bruker UK Ltd, Coventry for the loan of Microflex MALDI-ToF mass spectrometer and Dr Erika Tranfield and Dr Julie Green for technical support with Bruker MALDI-ToF.

